# Pipeline Evaluation of a State-of-the-Art AI Algorithm for Detection of Focal Cortical Dysplasia: Insights into Potential Failure Sources

**DOI:** 10.64898/2026.01.13.26344036

**Authors:** Mateus A. Esmeraldo, Stefanie Chambers, Yanniklas Kravutske, Eduardo Pontes Reis, Gregor Kasprian, Ana Filipa Geraldo, Sergios Gatidis, Bruno P. Soares

## Abstract

**Purpose:** MELD Graph is a state-of-the-art artificial intelligence (AI) model for automated detection of focal cortical dysplasia (FCD), but its performance remains limited, highlighting the need to investigate which aspects of the pipeline affect its accuracy.

**Methods:** A retrospective failure-mode analysis of the MELD Graph pipeline was performed in 242 subjects, with model predictions and FreeSurfer segmentations reviewed to classify errors as segmentation-associated or algorithm-related. FCD imaging features salient to humans were quantified, with statistical associations examined for both MELD Graph detection and focal FreeSurfer segmentation failure.

**Results:** MELD Graph demonstrated overall performance similar to previously published non-harmonized results, achieving a sensitivity of 69%, specificity of 44%, and positive predictive value (PPV) of 75%. Focal FreeSurfer segmentation failures were associated with 21% of false negative patients, 25% of false positive clusters in patients, and 16% of false positive clusters in controls. Higher conspicuity on T1-weighted images was associated with MELD Graph detection, whereas greater conspicuity on T2-FLAIR images relative to T1 was associated with detection failure. Bottom-of-sulcus dysplasia (BOSD) and presence of transmantle sign were not associated with detection. Non-BOSD lesions, higher human conspicuity measures, and low T1 image quality were positively associated with focal FreeSurfer segmentation failures.

**Conclusion:** FreeSurfer segmentation failures are a significant potential source of error in the MELD Graph pipeline. FCD imaging features salient to humans and image quality were also associated with variability in the algorithm performance. Robust cortical segmentation and stronger integration of T2-FLAIR imaging features may be beneficial for automated FCD detection tools.

## 1. Introduction

Epilepsy remains a major cause of neurological morbidity in both adults and children, with an estimated 0.6% of the global population actively affected[1]. Early surgical treatment can be curative (ILAE Class 1) in up to 70% of cases where it is caused by focal cortical dysplasia (FCD), one of the most common structural etiologies of drug-resistant epilepsy [2–4]. Therefore, early detection and referral for multidisciplinary surgical evaluation is a priority in the management of patients with drug-resistant epilepsy[5]. However, FCD is often difficult to identify on magnetic resonance imaging (MRI), as it may present with subtle or ambiguous imaging features. In a recently published benchmark study evaluating human performance in FCD detection on MRI examinations, expert detection rate was only 49%[6].

Lack of FCD detection contributes to substantial delay in treatment, with approximately 60% of patients with drug-resistant epilepsy not receiving appropriate surgical treatment [7]. Longer epilepsy duration is consistently associated with poorer neurocognitive outcomes and early surgery can prevent and even reverse cognitive decline, with measurable gains in memory, speed, and academic skills[8].

Computer-assisted lesion detection on imaging studies is one of the main areas where artificial intelligence (AI) could meaningfully advance patient care. Recent advances in AI have led to a growing number of models for automated FCD detection in MRI, reaching 41 AI-based studies in recent systematic review and meta-analysis on the topic [9]. However, AI models often face challenges in translating into routine clinical practice, particularly due to their limited performance on heterogeneous real-world data and lack or interpretability [3]. A recently developed open source and interpretable algorithm using graph neural networks (MELD Graph) has emerged as a state-of-the-art tool for FCD detection [2]. It shows strong potential for generalizability, having been trained and validated on heterogeneous data from 23 epilepsy centers worldwide and across all types of FCD.

Our experience with testing MELD Graph on real-world data shows a remarkable capability to detect very subtle FCDs that are frequently not detected by human readers. In contrast, during the quality control (QC)[10] of MELD Graph outputs in cases where the model prediction was incorrect, we noticed that FreeSurfer cortical segmentation failures at the FCD site are common, and these errors appeared to account for a substantial proportion of both false-positive controls and false-negative predictions in patients. Despite the important role of FreeSurfer in the MELD Graph pipeline, the impact of such segmentation failures has not been adequately addressed in the existing literature.

We also noticed that, when compared with expert human evaluation, the algorithm still fails to detect lesions that are readily identifiable to human experts, especially when the FCD is conspicuous on T2-FLAIR-weighted images but less apparent on T1-weighted images.

Therefore, the main goal of this study is to conduct a failure-mode analysis of the current MELD Graph pipeline. We aim to quantify the frequency of FreeSurfer segmentation failures, and to establish an association between imaging features and pipeline failures.

## 2. Methods

### 2.1. Datasets and Participants

Two publicly available cohorts were analyzed: the Bonn Open Presurgery MRI Dataset of people with epilepsy and FCD type II[11], and the Imaging Database for Epilepsy and Surgery (IDEAS)[12]. The Bonn dataset contributed 85 patients with FCD type II and 85 controls. The IDEAS dataset included 51 patients with postoperative histopathological confirmation of FCD and 100 controls.

In the Bonn cohort, one patient was removed due to persistent failure of the FreeSurfer cortical surface reconstruction. In the IDEAS cohort, 15 patients were excluded because their lesions were located outside the neocortex, predominantly within the hippocampus, which falls beyond the intended scope of MELD Graph[2].

All 85 Bonn controls were retained, and 36 IDEAS controls were included to match the number of IDEAS patients. The resulting pre–quality-control cohort consisted of 120 patients and 121 controls (241 subjects total).

### 2.2. Analytical Pipeline

This study was a retrospective cross-cohort failure-mode analysis. First, MELD Graph detection performance was assessed in both the full cohort and in the final cohort after automated QC, which was used in the original MELD Graph study design and is expected to identify subjects with widespread segmentation errors [2].

Second, a systematic manual review of MELD Graph predictions and FreeSurfer-derived cortical surfaces was performed to classify errors as segmentation-associated or algorithm related. Lesion-level human-salient imaging features were characterized using a framework that included bottom-of-sulcus dysplasia (BOSD) morphology, the presence of a transmantle sign, and a standardized conspicuity scoring system applied separately to T1 and T2-FLAIR imaging. FreeSurfer segmentation quality at each lesion was evaluated using predefined criteria to determine whether the abnormal cortex had been appropriately included within the reconstructed cortical ribbon. Finally, statistical analyses were conducted to examine associations between imaging features, MELD Graph detection outcomes, FreeSurfer segmentation failures, and image quality.

An overview of the full methodological pipeline is shown in **Figure 1**.

**Figure 1.**
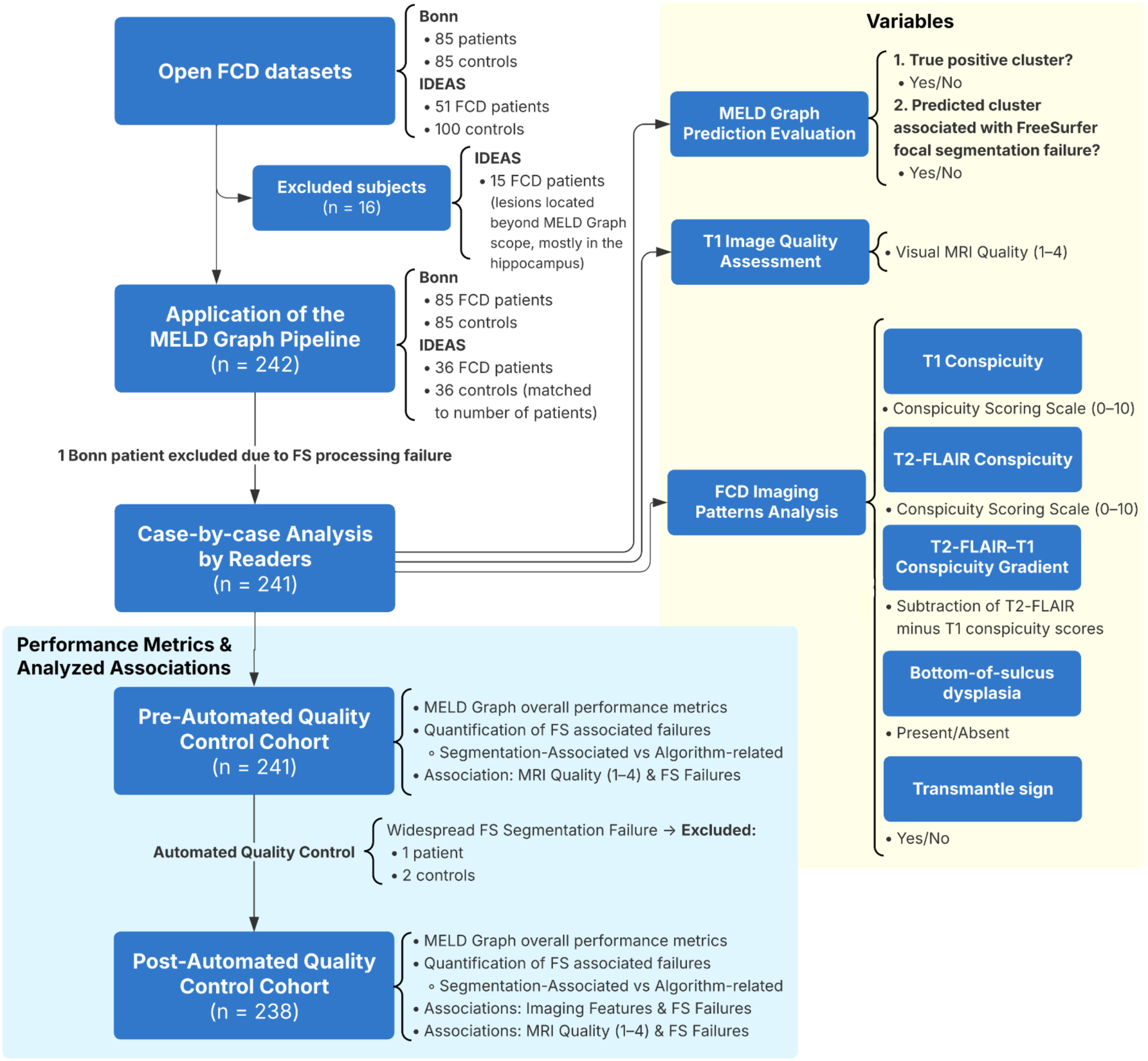
Overview of the study methodology. Abbreviations: FCD = focal cortical dysplasia; FS = FreeSurfer.

### 2.3. Preprocessing and Model Application

All image processing followed the procedures described in the original MELD Graph publication [2] and in the official MELD Graph documentation[10]. Cortical surfaces were reconstructed using FreeSurfer (v7.2.0) following the standard processing pipeline, and MELD Graph was applied with the latest publicly released model (version 2.2.2), using the default configuration [13,14]. To better simulate real-world clinical application, no retraining, parameter tuning, or data harmonization was performed.

Automated QC of surface-based features was conducted using the described MELD QC procedure[2,15], which identifies subjects with extreme structural or intensity values across cortical regions, typically resulting from imaging artefacts or widespread obvious FreeSurfer segmentation errors[15]. Performance metrics were calculated and reported both before and after automated QC. The ground truth lesion masks used in all analyses were the reference masks provided within each of the two open datasets[11,12].

### 2.4. Evaluation Metrics

Sensitivity, specificity, and positive predictive value (PPV) were defined and calculated as described in the MELD Graph supplementary material[2]. Each metric was computed across two stages of analysis: in the full cohort before automated QC, and in the final cohort after automated QC.

### 2.5. MELD Graph Pipeline Failure-Mode Analysis

All cases were reviewed in Freeview [13] by overlaying the MELD Graph predictions along with the pial and white matter surfaces onto the 3D T1-weighted images and, when available, onto the T2-FLAIR images as well[10]. This review was performed by a pediatric neuroradiologist (B.P.S.) with 20 years of neuroradiology experience and by a radiologist and postdoctoral research fellow in neuroradiology (M.A.E.). Discrepant interpretations were resolved in consensus including evaluation with an independent neuroradiologist (E.P.R.).

#### 2.5.1. Evaluation of FreeSurfer Segmentation Quality and Classification of MELD Graph Pipeline Errors

For each case, the accuracy of the white matter and pial surface reconstructions was assessed to determine whether the lesion was correctly included within the cortical ribbon (i.e., the region between the white matter and pial boundaries). Evaluation followed MELD Graph documentation guidelines[10]. A focal FreeSurfer segmentation failure was recorded when either of the following criteria was met:

- More than 5 mm of brain tissue was excluded from the region between the true pial surface and the nearest pial surface reconstruction (**Figure 2**). This relatively high threshold of 5 mm was chosen because smaller inaccuracies are common when using FreeSurfer[16].
- In the FCD patients, 50 percent or more of the lesion volume was excluded from the cortical segmentation.

**Figure 2.**
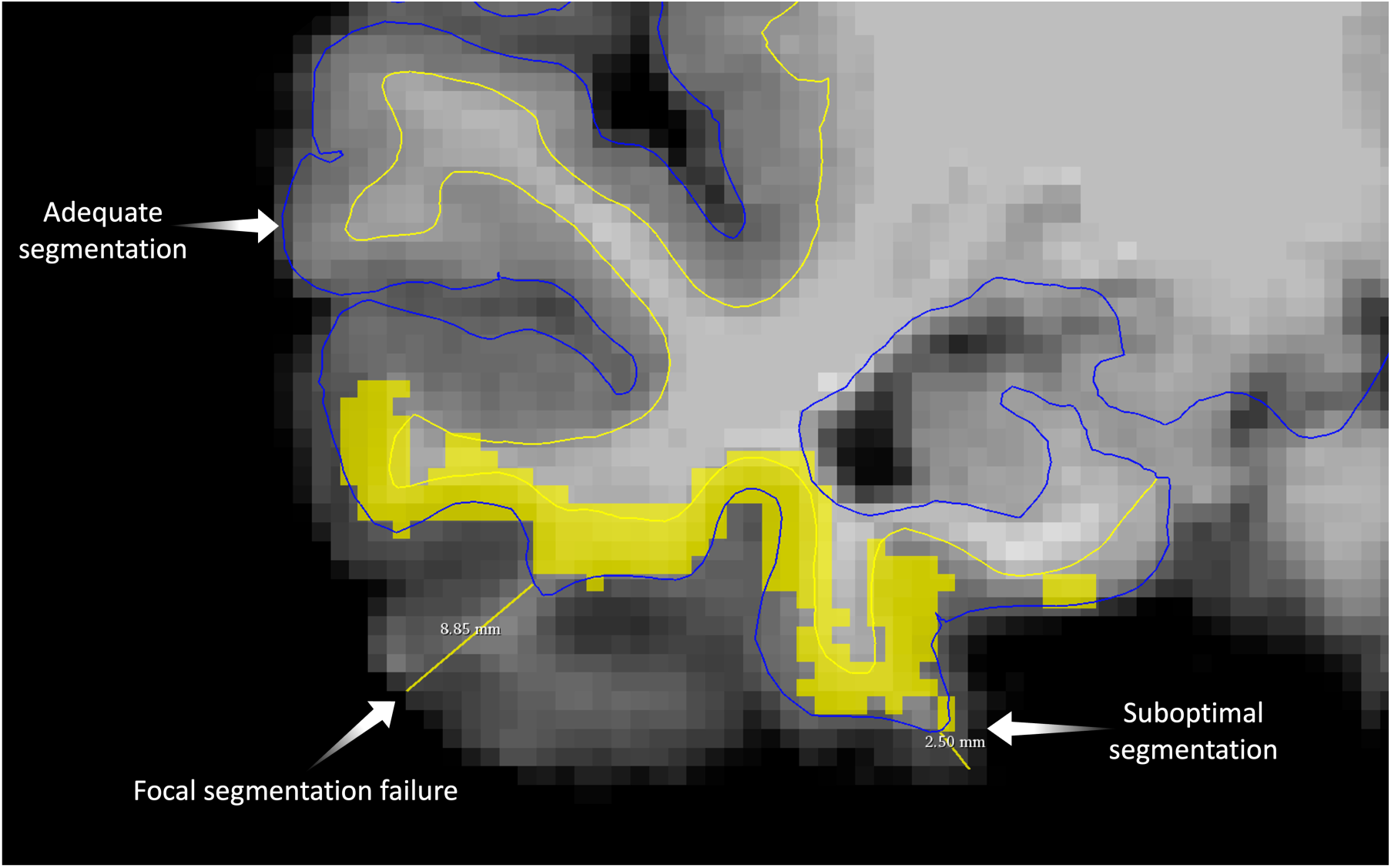
Illustration of manual quality assessment of FreeSurfer-derived cortical segmentation. The white matter (blue) and pial (yellow) surface reconstructions are shown over the T1-weighted image in Freeview[13]. Regions of adequate segmentation demonstrate close adherence of the surfaces to the true anatomical boundaries, whereas suboptimal segmentation shows deviations that fall within the predefined margin of error of less than 5 mm. A focal segmentation failure is also shown, in which more than 5 mm of cortical tissue is erroneously excluded from the region between the white matter and pial surfaces (measured as 8.85 mm in this control case). The yellow squares indicate the overlay corresponding to a false-positive MELD Graph prediction.

Following this segmentation assessment, each case was documented as either passing or failing cortical segmentation. Among the incorrect MELD Graph predictions (false positives and false negatives in patients with FCD, and false positives in controls), each was classified as either a FreeSurfer-associated focal segmentation failure or an algorithm (MELD Graph) failure, the latter referring to cases in which cortical segmentation was adequate, but the model prediction was incorrect.

### 2.6. FCD Patterns Assessment

Each FCD was assessed according to the following standard definitions:

- **Bottom-of-sulcus dysplasia (BOSD):** a subtype of FCD type II where abnormalities are maximal at the bottom of a single sulcus, potentially reaching but not extending over the gyral crown, regardless of subcortical signal change[17].
- **Transmantle sign:** linear or triangular shaped extension of high T2-FLAIR white matter signal intensity extending towards the ventricle[18].
- **FCD conspicuity:** FCD conspicuity in T1-weighted and T2-FLAIR imaging was rated using an adapted lesion conspicuity scoring system, described below.

#### 2.6.1. FCD Conspicuity Scoring Scale and Image Quality Assessment

To quantify lesion visibility, we adapted a previously described conspicuity scale specifically for FCD evaluation[19]. The scoring range was expanded from 0 to 10 to capture the wide variability and complexity of FCD assessment [6,19]. Separate scores are assigned for T1-weighted and T2-FLAIR images.

Readers first attempted to localize the lesion without prior information using the original NIfTI images. After this blinded assessment, the ground truth lesion mask was revealed, and a conspicuity score was assigned (**Table 1**). Scores were determined by consensus between the two primary study readers (B.P.S. and M.A.E.), with discrepant cases resolved in consultation with an independent neuroradiologist (E.P.R.). For such cases, the final score was defined as the average of all three ratings.

**Table 1.** Focal Cortical Dysplasia (FCD) Conspicuity Scoring Scale. The scale ranges from 0 to 10, with higher scores indicating greater conspicuity. The descriptions provided correspond to key reference levels that orient scoring along the continuum; intermediate values represent proportional gradations between these defined levels. Abbreviations: FCD = Focal cortical dysplasia; EEG = electroencephalography; PET = positron emission tomography.

| Score | Description |
| --- | --- |
| 10 – Maximal conspicuity | The FCD is readily visible; the reader considers that even an untrained person with no medical training could identify it with minimal guidance. |
| 5 – Moderate conspicuity | The FCD would be detected by most radiologists using a standard search strategy and is readily identifiable to most neuroradiologists and epilepsy-imaging experts. |
| 1 – Minimal conspicuity | The FCD is extremely subtle, generally not detectable without multimodal information (e.g., EEG, PET) that restricts the search region. |
| 0 – Non-detectable | The FCD is considered non-identifiable on MRI if it cannot be detected by two neuroradiologists, allowing substitution of one reviewer by an epilepsy-imaging specialist, even when its precise location is disclosed. |

Image quality was assessed using the standardized 4-point visual quality control scale [20]. Since MELD Graph relies on FreeSurfer-derived cortical surface reconstruction, and FreeSurfer primarily uses T1-weighted images, we evaluated T1 image quality for its potential impact on FreeSurfer cortical segmentation failures. In patients, only segmentation failures not attributable to the FCD itself were included, since errors within the dysplastic cortex typically reflected lesion-related anatomic or signal abnormalities and were evaluated separately as FreeSurfer-associated focal segmentation failure, as previously described. In controls, all focal segmentation failures were included.

### 2.7. Statistical tests

All statistical analyses were performed to evaluate associations between FCD imaging features, MELD Graph FCD detection, and focal FreeSurfer segmentation failures. Continuous variables derived from the conspicuity scoring system were analyzed using point-biserial correlation to assess their relationship with binary outcomes, including MELD Graph detection (detected vs not detected) and the presence of focal FreeSurfer segmentation failure (failure vs no failure). Effect sizes for these correlations were reported as Pearson r values with corresponding t statistics and p values.

Categorical imaging features, including BOSD morphology and the presence of a transmantle sign, were evaluated using chi-square tests of independence. Effect sizes for chi-square tests were expressed using Phi (φ). When a chi-square test demonstrated a statistically significant association, relative risk (RR) and 95 percent confidence intervals were calculated to quantify the magnitude of the effect.

The association between T1 image quality and the occurrence of focal FreeSurfer segmentation failure was assessed using Spearman rank correlation. T1 image quality was treated as an ordinal variable on the standardized 1 to 4 visual quality scale[20], and segmentation failure was coded as a binary variable (1 = present, 0 = absent).

All tests were two-tailed. Statistical analyses were performed in Python (version 3.13.5) using the pandas, NumPy, and SciPy libraries.

## 3. Results

All individual-level data, including imaging features, segmentation outcomes, and MELD Graph predictions of FCD cases and controls are provided in **Supplementary Tables 1** and **2**, respectively. All counts, including totals of false negatives, false positives, true positive clusters, and FreeSurfer segmentation failures are detailed in **Table 2**.

**Table 2.**
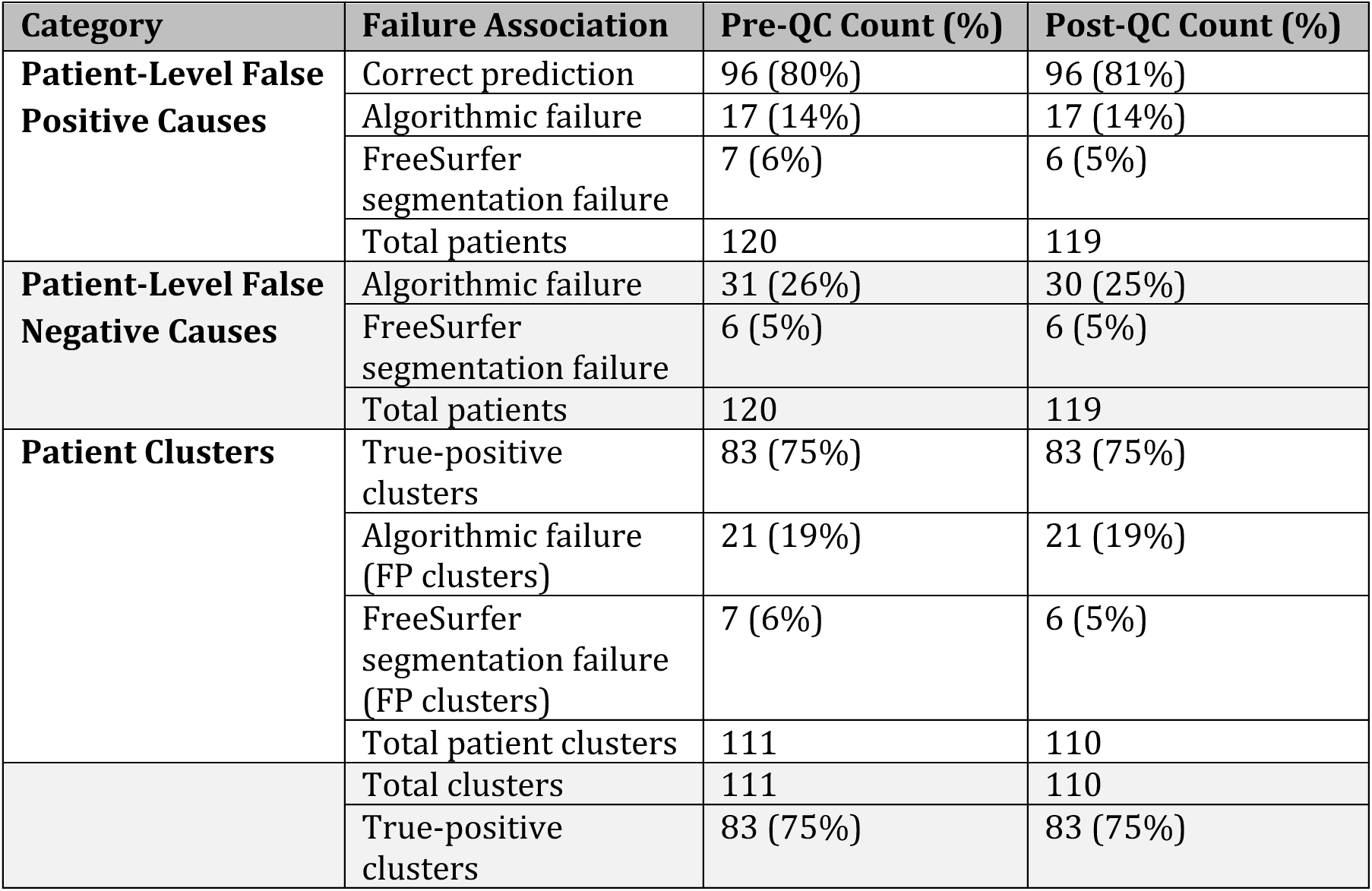
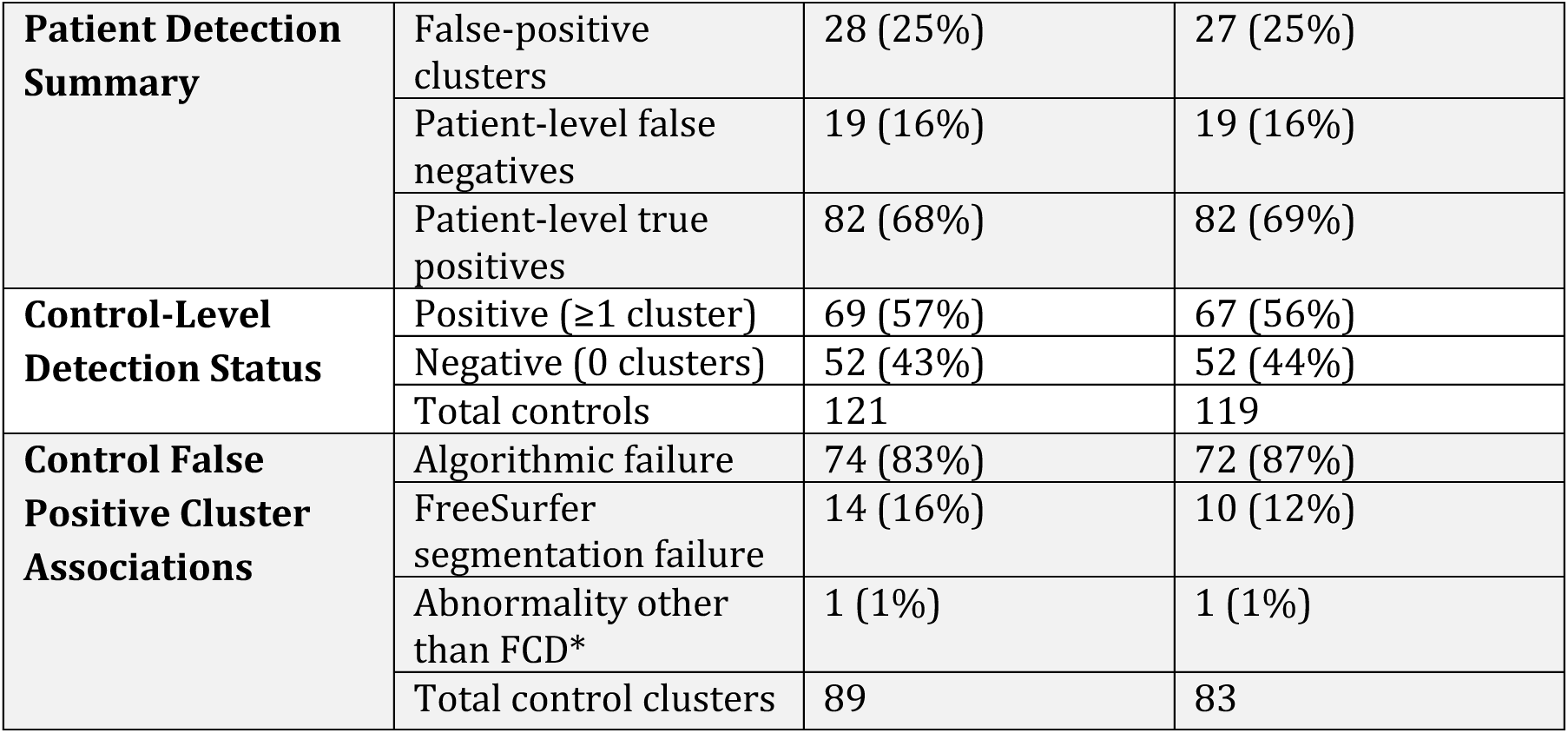
Detection Outcomes and Association with FreeSurfer Segmentation Failure in Pre- and Post-Automated Quality Control. Summary of the distribution of MELD Graph detection errors and their association with FreeSurfer focal segmentation failures in the pre- and post-automated QC cohorts. Abbreviations: QC = quality control; FS = FreeSurfer; FP = false positive; FN = false negative. *Although sub-00023 was classified as a control in the open dataset[11], the subject in fact had a true ischemic lesion in the banks of the left superior temporal sulcus. As MELD Graph accurately detected this abnormality, and given that control subjects are presumed to be lesion-free, this instance was not categorized as a MELD Graph detection failure.

### 3.1. Overall Performance and Association with FreeSurfer Segmentation Errors

Across the full cohort of 120 patients and 121 controls (241 subjects), MELD Graph achieved a sensitivity of 68%, specificity of 43%, and PPV of 75%. After automated QC, three subjects with widespread FreeSurfer segmentation errors were removed (one patient and two controls), and performance remained similar (sensitivity 69%, specificity 44%, PPV 75%). An example of a correct MELD Graph prediction is presented in **Figure 3**.

**Figure 3.**
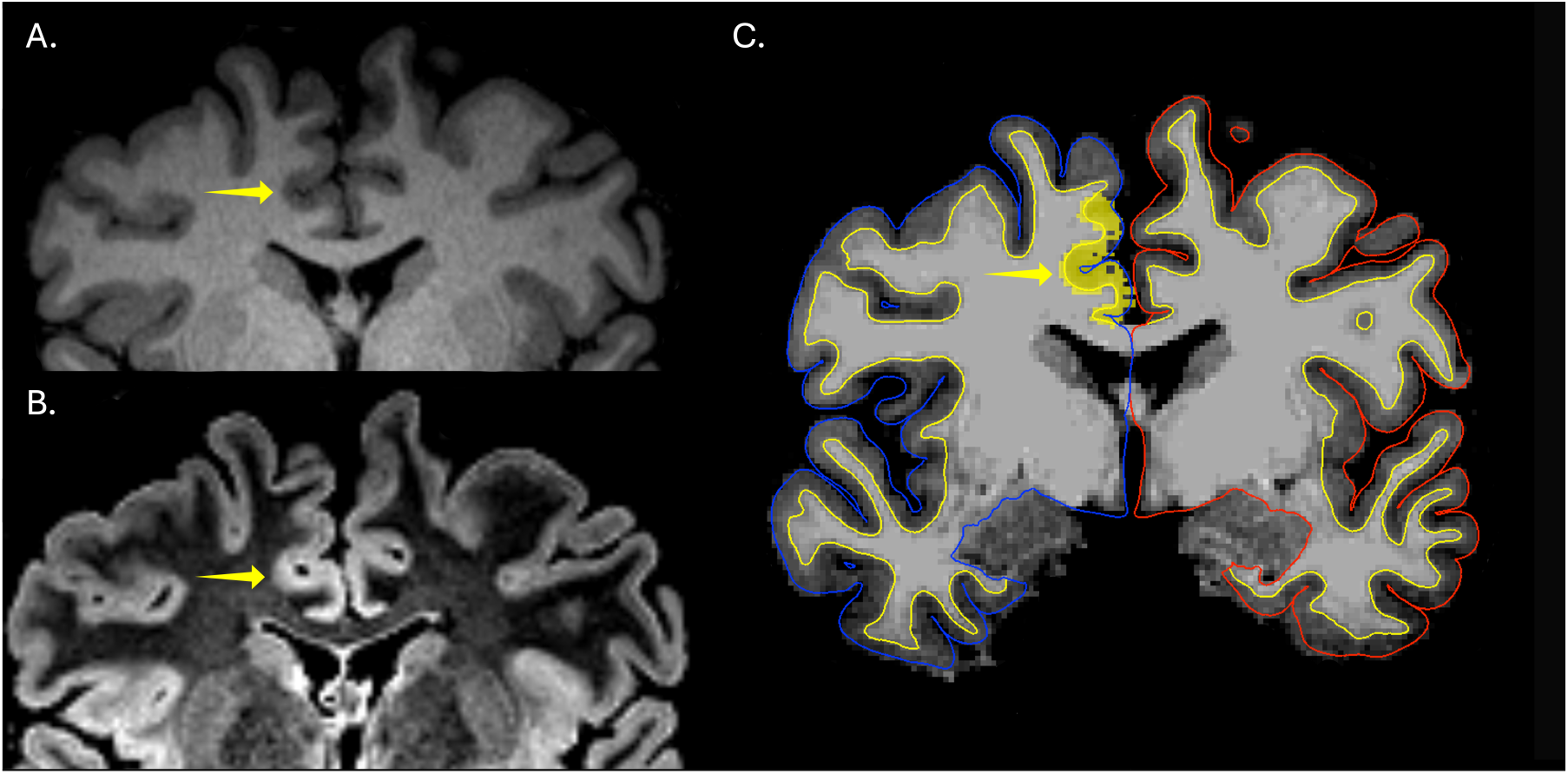
Detection of a minimally conspicuous focal cortical dysplasia (FCD) by MELD Graph. Coronal T1-weighted (A) and T2-FLAIR (B) brain MRI images demonstrating an extremely subtle right frontal paramedian FCD (yellow arrows). The lesion was rated as minimally conspicuous by expert human readers and was considered generally nondetectable without multimodal information to constrain the search region. **(C)** MELD Graph correctly identified the FCD, yielding a true-positive prediction despite its subtle imaging appearance to human readers.

In the pre-automated QC cohort, focal FreeSurfer segmentation failures in the FCD were associated with 4 of 19 false negatives in patients (21%), 7 of 28 false positive clusters in patients (25%), and 14 of 88 false positive clusters in controls (16%). After automated QC, focal segmentation failures were associated with 4 of 19 false negatives in patients (21%), 6 of 27 false positive clusters in patients (22%), and 10 of 82 false positive clusters in controls (12%). **Figure 4** exemplify a false-negative case associated with a focal segmentation failure, whereas **Figure 5** demonstrates a case with adequate segmentation in which MELD Graph nonetheless failed to detect the lesion.

**Figure 4.**
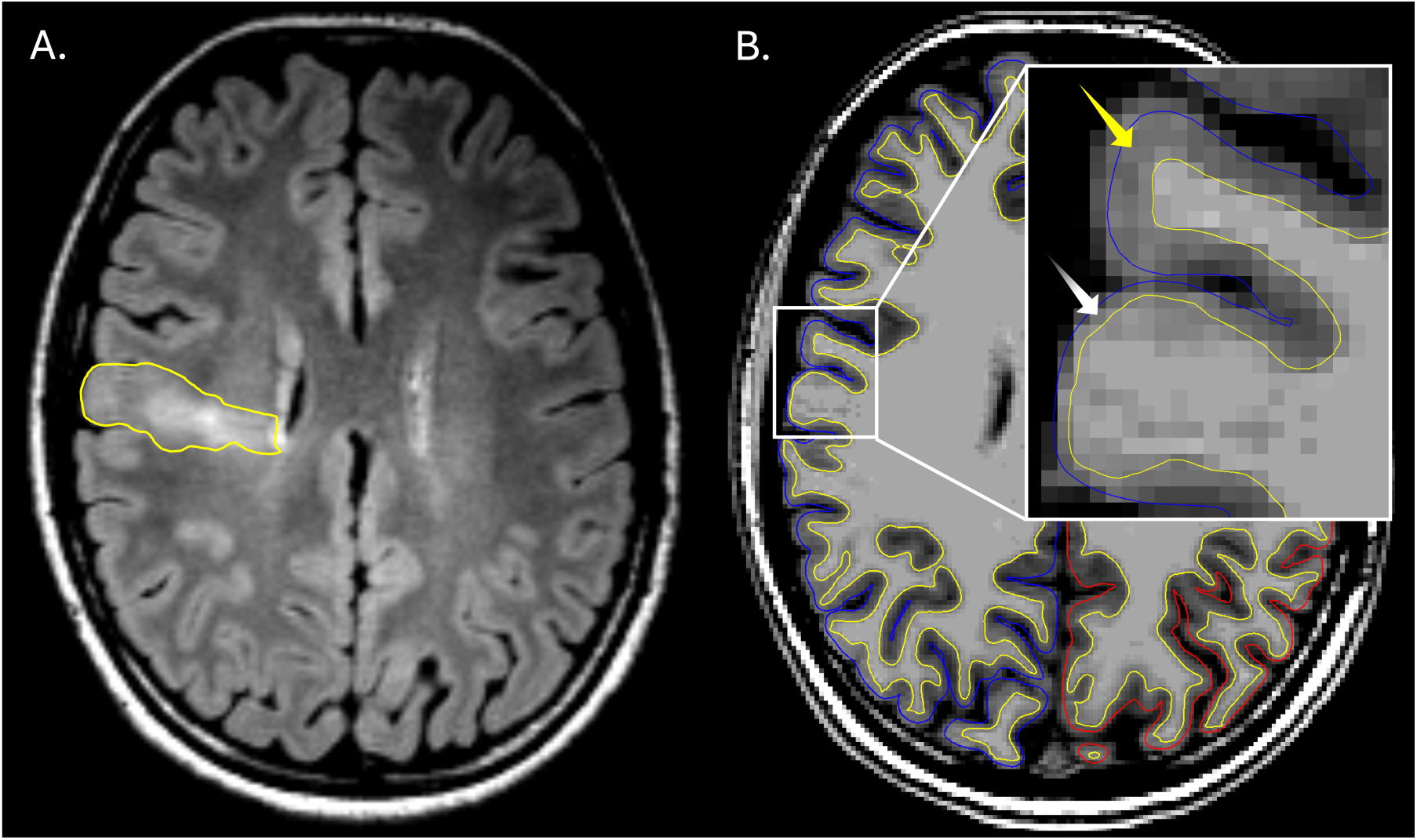
False negative prediction associated with FreeSurfer focal cortical segmentation failure. **(A)** Axial T2-FLAIR image demonstrating a right frontal FCD, outlined by the yellow contour, rated as maximally conspicuous by human readers. **(B)** Axial T1-weighted image with FreeSurfer-derived white matter (yellow line) and pial (blue line in the right hemisphere) surface reconstructions overlaid in Freeview[10,13]. The inset highlights two key segmentation examples: the yellow arrow indicates an area of cortical segmentation that is considered acceptable, while the white arrow demonstrates a substantial segmentation failure in which most of the lesion lies outside the cortical segmentation. This error resulted from overexpansion of the white matter mask toward the pial surface, causing most of the dysplastic cortex to be mislabeled as white matter rather than cortex. This FCD, although readily identifiable to human readers, was not detected by MELD Graph, likely due primarily to this segmentation error.

**Figure 5.**
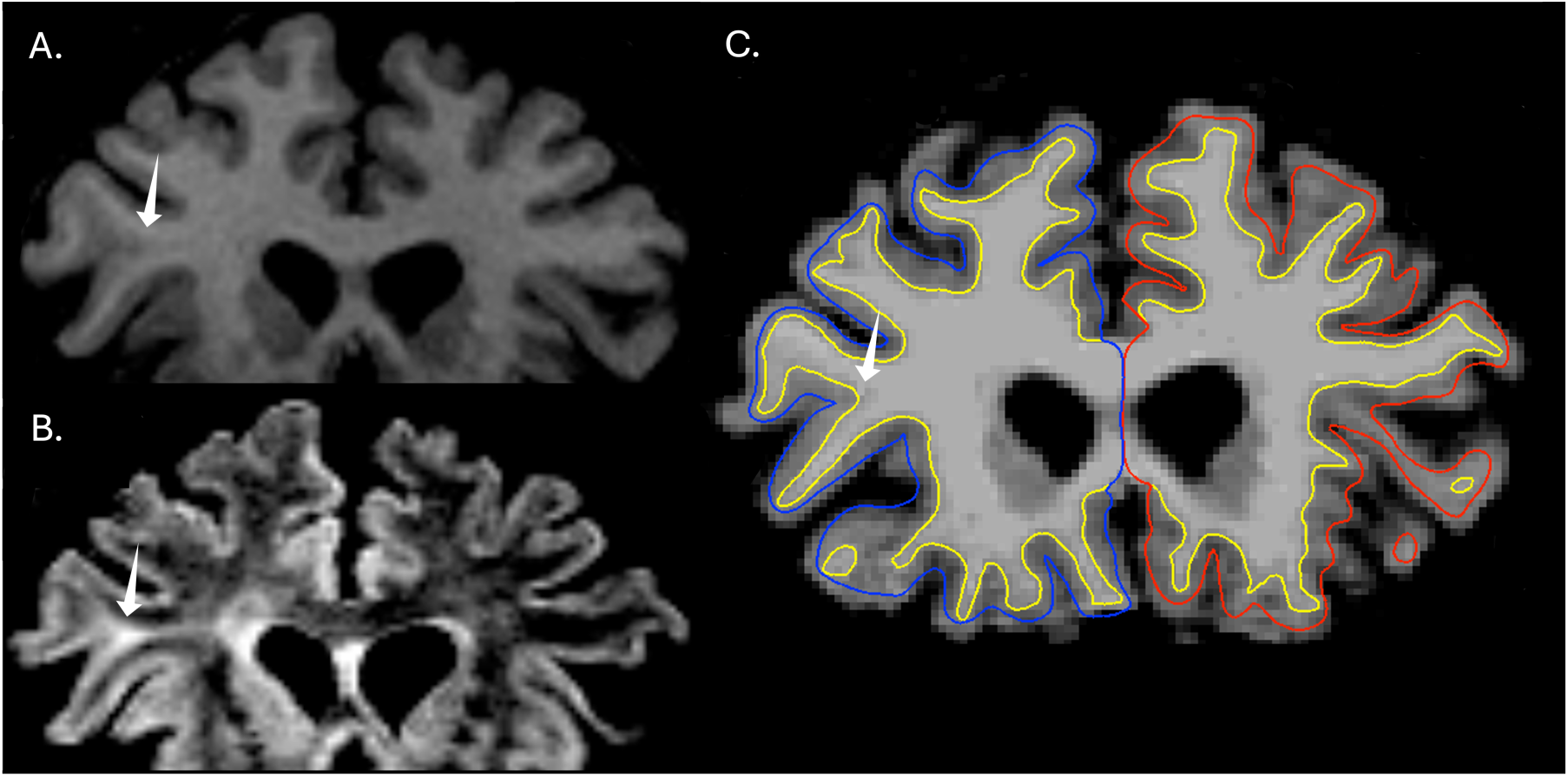
Focal cortical dysplasia (FCD) conspicuous in T2-FLAIR images but not detected by MELD Graph. Coronal T1-weighted **(A)** and T2-FLAIR (B) brain MRI images demonstrating a right frontal FCD with a transmantle signs (white arrows). The lesion was particularly evident on the T2-FLAIR sequence but less apparent on T1-weighted imaging. **(C)** MELD Graph failed to detect the abnormality even with adequate cortical segmentation by FreeSurfer, yielding a false-negative prediction despite its clear appearance to human specialists.

### 3.2. Correlations between FCD Imaging Features and MELD Graph Detection

We found a statistically significant positive correlation between T1WI conspicuity and lesion detection (r = 0.35, p = 0.0001; n = 119), indicating that FCD more visible on T1-weighted images were more likely to be detected. Conversely, T2-FLAIR conspicuity demonstrated a weaker positive correlation trend (r = 0.21, p = 0.056; n = 83), which did not reach statistical significance.

Assessment of the T2-FLAIR–T1 conspicuity gradient revealed a significant negative correlation with MELD Graph detection (r = –0.24, p = 0.036; n = 76), indicating that FCDs with higher conspicuity on T2-FLAIR relative to T1 were less likely to be detected.

There were no statistically significant associations between FCD morphology (φ = 0.08, p = 0.38; n = 119), and presence or absence of transmantle sign (φ = 0.09, p = 0.42; n = 83) and MELD Graph performance.

### 3.3. FCD Imaging Features and FreeSurfer Focal Segmentation Failures

Higher conspicuity of the FCD on both T1 and T2-FLAIR images was significantly positively correlated with the likelihood of segmentation failure (T1: r = 0.33, p = 0.0002, n = 119; T2-FLAIR: r = 0.31, p = 0.00397, n = 83). The T2-FLAIR–T1 conspicuity gradient demonstrated a similar positive correlation (r = 0.30, p = 0.00835; n = 76), suggesting that lesions with disproportionately higher T2-FLAIR conspicuity are more likely to be incompletely segmented. Moreover, FCD morphology (BOSD vs non-BOSD) showed a significant association with segmentation failure (φ = 0.31, p = 0.00059; n = 119), with non-BOSD lesions exhibiting a markedly increased risk of segmentation failure (RR = 7.82; 95% CI: 1.85–32.7). We found no statistically significant association between presence or absence of the transmantle sign and segmentation failure (φ = 0.03, p = 0.76; n = 83).

### 3.4. Image Quality Assessment and FreeSurfer Segmentation Failure

T1 image quality showed a significant negative correlation with the occurrence of FreeSurfer segmentation failures. Before automated QC, lower image quality was associated with a higher likelihood of segmentation failure (ρ = –0.19, p = 0.0039). This association remained significant, although slightly attenuated, after exclusion of the three cases removed by automated QC for widespread FreeSurfer segmentation errors (ρ = -0.16, p = 0.016), indicating that lower T1 image quality was associated with a higher likelihood of focal FreeSurfer segmentation failure.

A summary of the results is provided in **Table 3**.

**Table 3.**
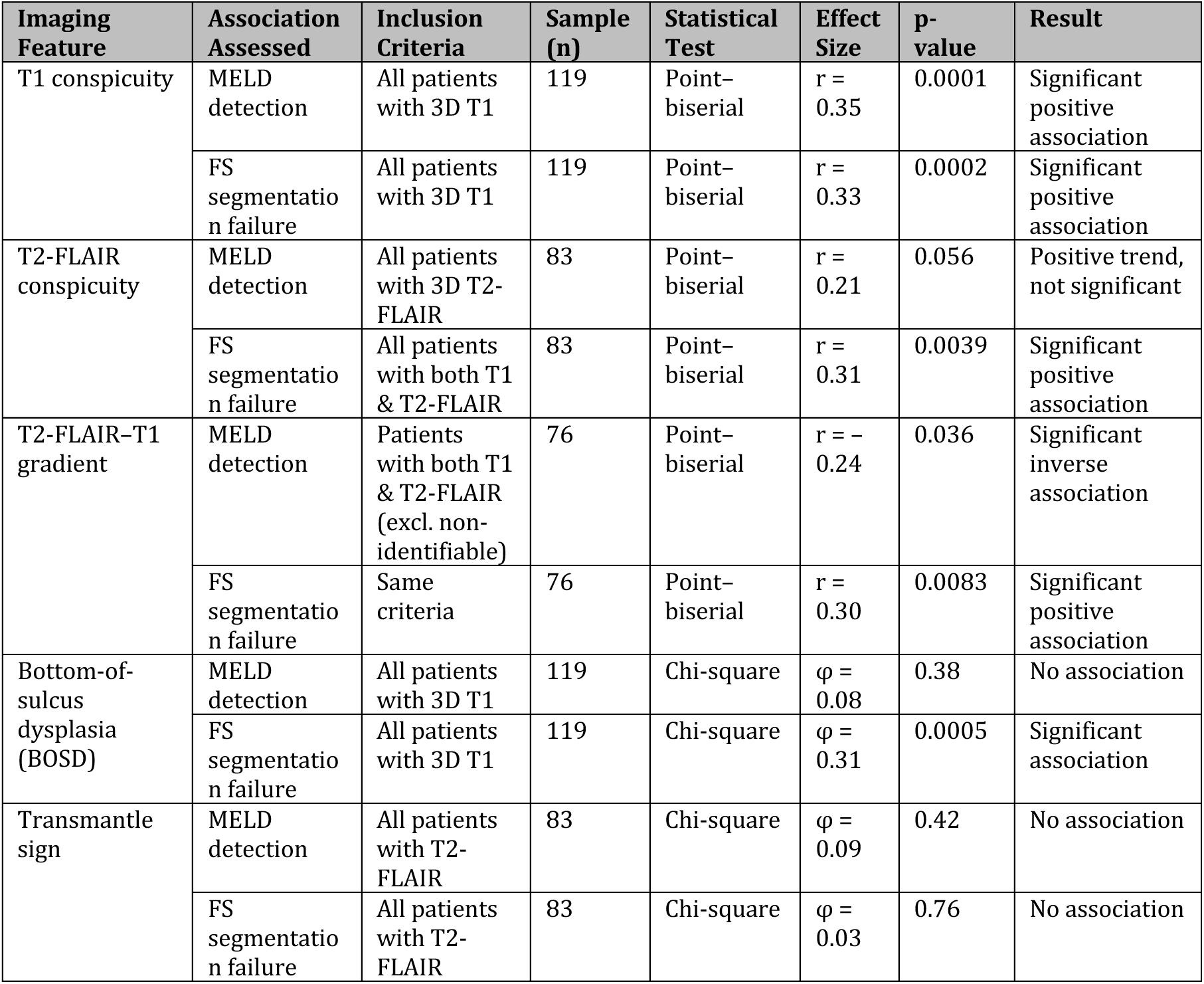
Association Between MELD Graph detection, Focal Cortical Dysplasia Imaging Features and FreeSurfer Focal Segmentation Failures. Summary of associations between human-salient FCD imaging features, MELD Graph lesion detection, and focal FreeSurfer cortical segmentation failure. Abbreviations: FCD = focal cortical dysplasia; FS = FreeSurfer; FLAIR = fluid-attenuated inversion recovery; T1WI = T1-weighted imaging; r = Pearson point–biserial correlation coefficient; φ = Phi coefficient.

## 4. Discussion

In this independent evaluation of MELD Graph applied to two publicly available, heterogeneous cohorts of brain MRI exams of subjects with FCD and healthy controls, performance showed a sensitivity of 69%, specificity of 44%, and a PPV 75%, which was broadly consistent with that reported in the original multicenter study under non-harmonized conditions, which demonstrated a sensitivity of 70%, specificity of 39%, and a PPV of 70% in the independent test cohort[2].

Our results show that focal FreeSurfer segmentation failures are associated with a substantial proportion of MELD Graph errors (12-25% of incorrect predictions). Although automated quality control was used in the original MELD Graph description[2,15], it is designed to remove only subjects with obvious and widespread artifacts or FreeSurfer reconstruction failures. Indeed, it eliminated only three of 242 subjects in our study and only 14 subjects in the original MELD report, which included 618 patients[15]. Consequently, automated QC may create a false impression that segmentation errors have largely been excluded, when in reality a substantial number of focal segmentation failures persist, such as demonstrated in our results. This illustrates that segmentation problems represent a significant drawback of the algorithm performance, and that correcting such focal errors represents an opportunity for potentially improving MELD Graph diagnostic accuracy.

Through a root-cause analysis of cases with segmentation failure, we have noticed that when the reconstructed cortical ribbon was distorted, the region labeled as cortex was often composed of white matter or was misaligned with the true cortical boundaries, producing thickness, curvature, sulcal depth, and intensity values that did not reflect the underlying anatomy, as illustrated in **Figure 2** and **Figure 4**. In these situations, MELD Graph was effectively operating on corrupted input features. This observation is even more relevant when considering that all training of MELD Graph was performed on cortical surfaces reconstructed with FreeSurfer[2,15], which potentially introduced a substantial number of inadequately segmented cases into the training data. These findings indicate that addressing focal FreeSurfer segmentation failures and correcting them before training can provide the model with cleaner representations of lesion-related cortical features, representing a realistic and relatively low-effort opportunity to enhance MELD Graph performance. However, considering that these focal errors required meticulous case-by-case inspection to be identified, this level of manual review poses a substantial barrier to clinical translation for the MELD Graph pipeline, since it is time consuming and unfamiliar to most clinicians[3].

We have also noticed a consistent association between FCD lesion appearance and segmentation failures, which has the potential to guide future AI algorithmic improvement. Lesions that were highly conspicuous to human readers were also more likely to be poorly segmented by FreeSurfer, reflecting the fact that the same anatomical distortions that make FCDs evident to human visual inspection can destabilize automated surface reconstruction. In addition, FreeSurfer fundamentally requires proper gray–white matter contrast to accurately reconstruct the cortical ribbon, yet gray–white matter blurring is the most frequent imaging hallmarks of FCD[21,22], making it particularly unsuitable in the FCD context. Subsequent quantitative analysis of FCD T1WI conspicuity, T2-FLAIR conspicuity, and their gradient brought additional insights into potential causes for differences between human and MELD Graph performance, and provided a framework for human–algorithm comparisons. Indeed, in our sample, T2-FLAIR–T1 conspicuity gradient captured cases in which lesions were more visible on T2-FLAIR relative to T1, a pattern that human readers typically identify with relative ease. This is in line with the fact that T2-FLAIR is widely regarded as the most informative sequence for FCD detection by human readers[22]. However, according to our results T2-FLAIR–T1 conspicuity gradient was also significantly associated with both MELD Graph non-detection and a higher likelihood of focal segmentation failure. It is remarkable that MELD Graph achieves its performance relying mainly on T1-weighted imaging[2], given that the conspicuity scores were lower on T1 than on T2-FLAIR in all our subjects. Nevertheless, this is not completely unexpected as MELD Graph considers T2-FLAIR as an optional sequence, and the addition of the latter yielded only a modest, non-significant increase in detection rate from 67.3% to 72.7% in the original study[2]. Taken together, these findings suggest that integrating T2-FLAIR more centrally into FCD detection pipelines may improve their performance.

Moreover, we have also found that BOSD (an often subtle FCD subtype commonly identified in patients with an initial “non-lesional” or “negative” brain MRI [17]) were much less likely to generate focal FreeSurfer segmentation failures, indicating that the current MELD Graph pipeline handles their geometry effectively. However, according to our results they were not detected more frequently than non–BOSD lesions, meaning that reliable segmentation alone is not sufficient for consistent automated identification.

Conversely, lower T1WI image quality was significantly positively correlated with likelihood of focal cortical segmentation failures, underscoring the need for high-quality imaging acquisition of this sequence when evaluating patients with FCD. These findings reinforce the importance of referring patients with suspected FCD to centers with specialized epilepsy-imaging protocols, high-quality hardware and rigorous acquisition standards[5].

Although providing the first systematic quantitative and qualitative evaluation of the MELD Graph errors on FCD detection, this study is not without limitations. Segmentation failures were documented but not manually corrected, so the exact performance gain that would occur in the baseline algorithm if these errors were corrected cannot be determined. A future study focused specifically on segmentation failures could incorporate targeted correction or adopt more modern segmentation methods, such as approaches based on deep learning architectures including nnU-Net, SegResNet, UNETR, SwinUNETR, and U-Mamba_Bot model, which achieved high overall accuracy in delineation of gray matter structures [23]. On the other side, the binarization of features such as BOSD morphology and the transmantle sign oversimplify the heterogeneity of FCD imaging patterns. For example, the transmantle sign was coded as present or absent, grouping together lesions with widely different degrees and extension of subcortical T2-FLAIR abnormality. Some cases showed a clear subcortical T2 abnormal signal, such as the case depicted in **Figure 2**, while others were extremely subtle and only detectable with careful optimization of grayscale display settings. It is likely that only more evident transmantle T2-FLAIR hyperintensities would meaningfully affect FreeSurfer segmentation or MELD Graph detection, but the sample size was not large enough to aim to distinguish between these different levels of abnormal T2-FLAIR signal. Finally, the conspicuity scoring system used in this study has not yet been formally validated. Although the scale was adapted from existing frameworks[6,19], a dedicated validation study is needed to establish a reliable measure of human-perceived FCD conspicuity. Such a tool could help guide the development of future algorithms by quantifying imaging features that are particularly salient to expert readers and integrating them with computational approaches.

## 5. Conclusion

Our failure-mode analysis shows that FreeSurfer segmentation failures are a relevant source of error in the MELD Graph automated FCD detection pipeline. Lesion conspicuity, specific FCD imaging patterns, and image quality were also associated with performance variability and segmentation failures, highlighting opportunities to improve image processing and to integrate T2-FLAIR more centrally into the model architecture. Those findings highlight the importance of robust cortical segmentation for reliable automated FCD detection and provide insight into which expert human-salient FCD imaging features may inform the next generation of AI tools.

## Data Availability

All data used in this study were obtained from two publicly available open datasets: the Bonn Open Presurgery MRI Dataset of people with epilepsy and FCD type II[11] and the Imaging Database for Epilepsy and Surgery (IDEAS)[12]. All processed tables and code used in the analysis are available from the corresponding author upon request.

https://openneuro.org/datasets/ds004199/versions/1.0.2

https://openneuro.org/datasets/ds005602/versions/1.0.0

## IRB / Ethics Approval Statement

This study used only fully de-identified, publicly accessible data and did not involve prospective data collection or interaction with human participants[11,12]. According to institutional and national guidelines, Institutional Review Board (IRB) approval was not required.

## Ethics Approval and Consent to Participate

Not applicable. All imaging data were previously collected under ethical approvals by the original dataset providers and released in accordance with their respective consent procedures [11,12].

## Patient Consent Statement

Not applicable. This study analyzed de-identified, publicly released data[11,12] and involved no direct patient contact.

## Permission to Reproduce Material From Other Sources

No copyrighted or previously published material requiring permission for reproduction was used in this manuscript. All figures and analyses are original and based on publicly available datasets[11,12].

## Ethical Publication Statement

We confirm that we have read the *Brain Informatics* journal’s position on ethical publication and affirm that this manuscript complies with those guidelines.

## Clinical Trial Registration

Not applicable. This study is a retrospective analysis of previously acquired open-source imaging datasets and does not constitute a clinical trial.

## Author Contributions

Study conception and design: M.A.E., B.P.S., S.C. Data curation, preprocessing, and imaging review: M.A.E., B.P.S., E.P.R. MELD Graph pipeline evaluation: M.A.E., B.P.S., S.C. Statistical analysis and interpretation of results: M.A.E., S.C., Y.K. Manuscript drafting and critical revision: M.A.E., B.P.S., A.F.G., G.K., S.G., S.C., Y.K. Final approval of the submitted version: All authors.

